## Supplement for "Social isolation is linked to declining grey matter structure and cognitive functions in the LIFE-Adult panel study"

The corresponding contrast matrix was [0 0 0 0 0 0 0 0 0 0 0 1].

**Fig. S1**

Directed acyclic graphs demonstrating the theoretical underpinnings of model 1 and 2.

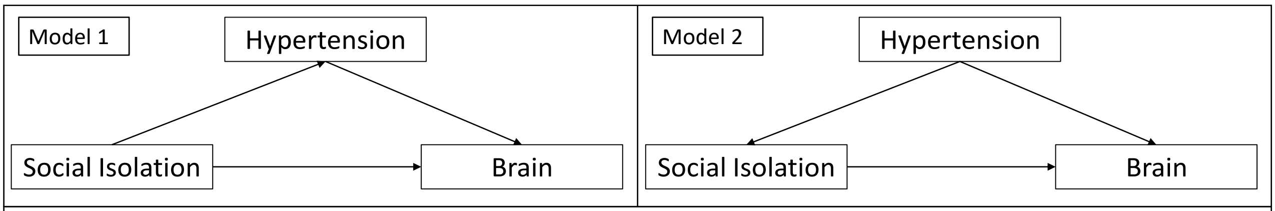


In model 1 the additional risk factors are assumed to be mediators and do not have to be controlled for. In model 2 they are assumed to be confounders. Therefore, they have to be controlled for.

#### Fig. S2

Simplified plot of the bivariate latent change score models

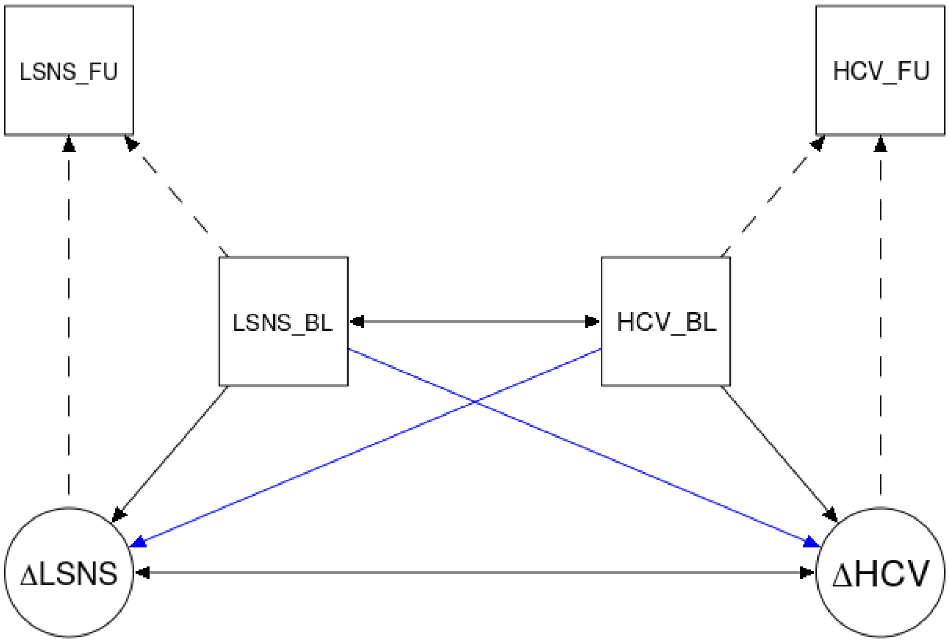


LSNS, Lubben Social Network Scale; HCV, hippocampal volume; BL, baseline; FU, follow-up; Δ, change in.

The blue arrows show our paths of interest.

**Fig. S3**

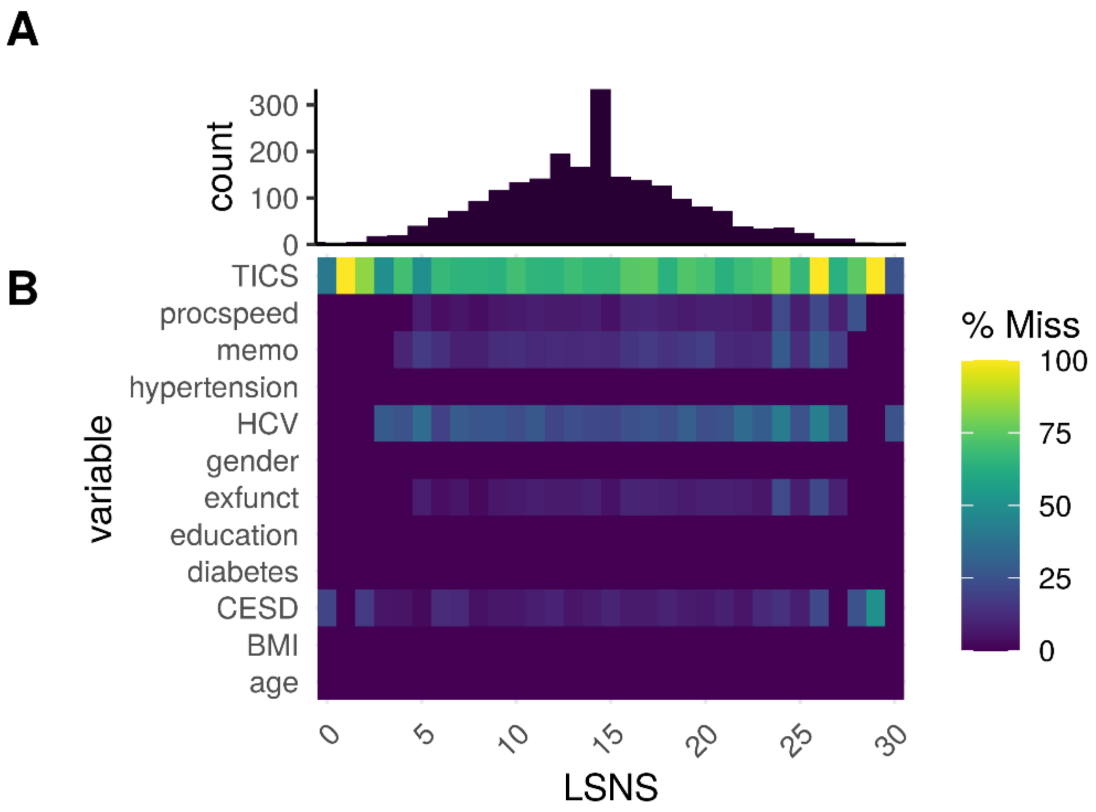


A) Histogram of LSNS scores by individual observation. B) Heatmap of proportional missingness of variables for different LSNS scores.

**Fig. S4**

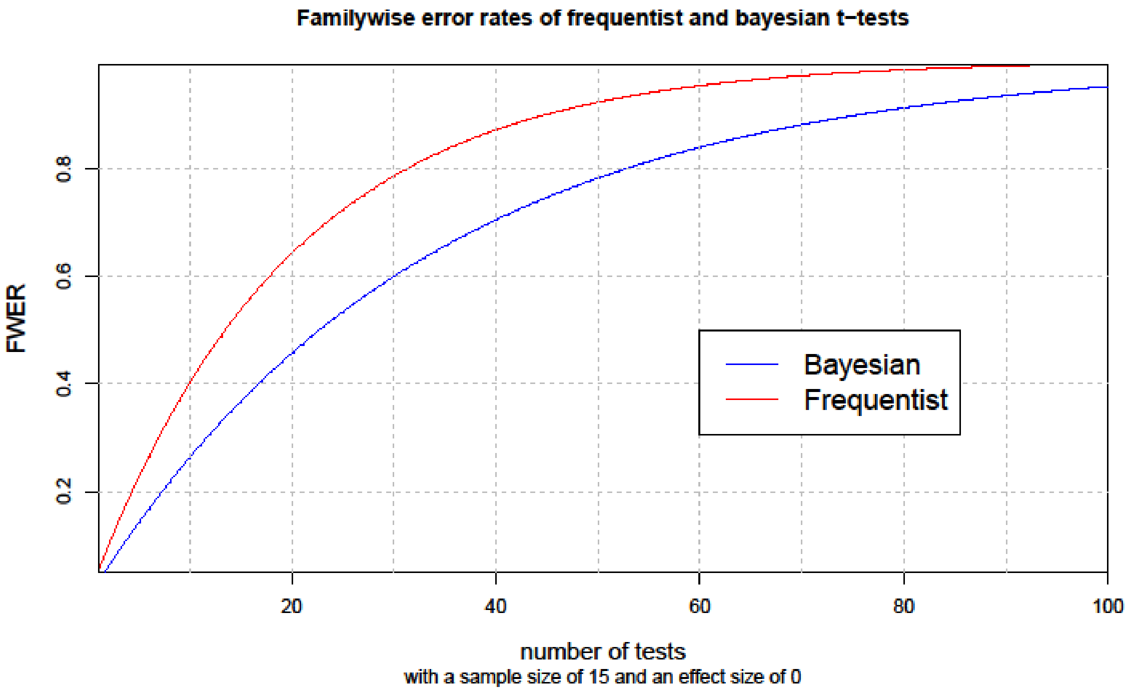
Familywise error rates of frequentist and bayesian t−tests
FWER
0.2
0.4
0.6
0.8
20
40
60
80
100
Bayesian
Frequentist
number of tests
with a sample size of 15 and an effect size of 0

**Fig. S5**

Histogram of BFs with randomly simulated values for our predictors of interest.

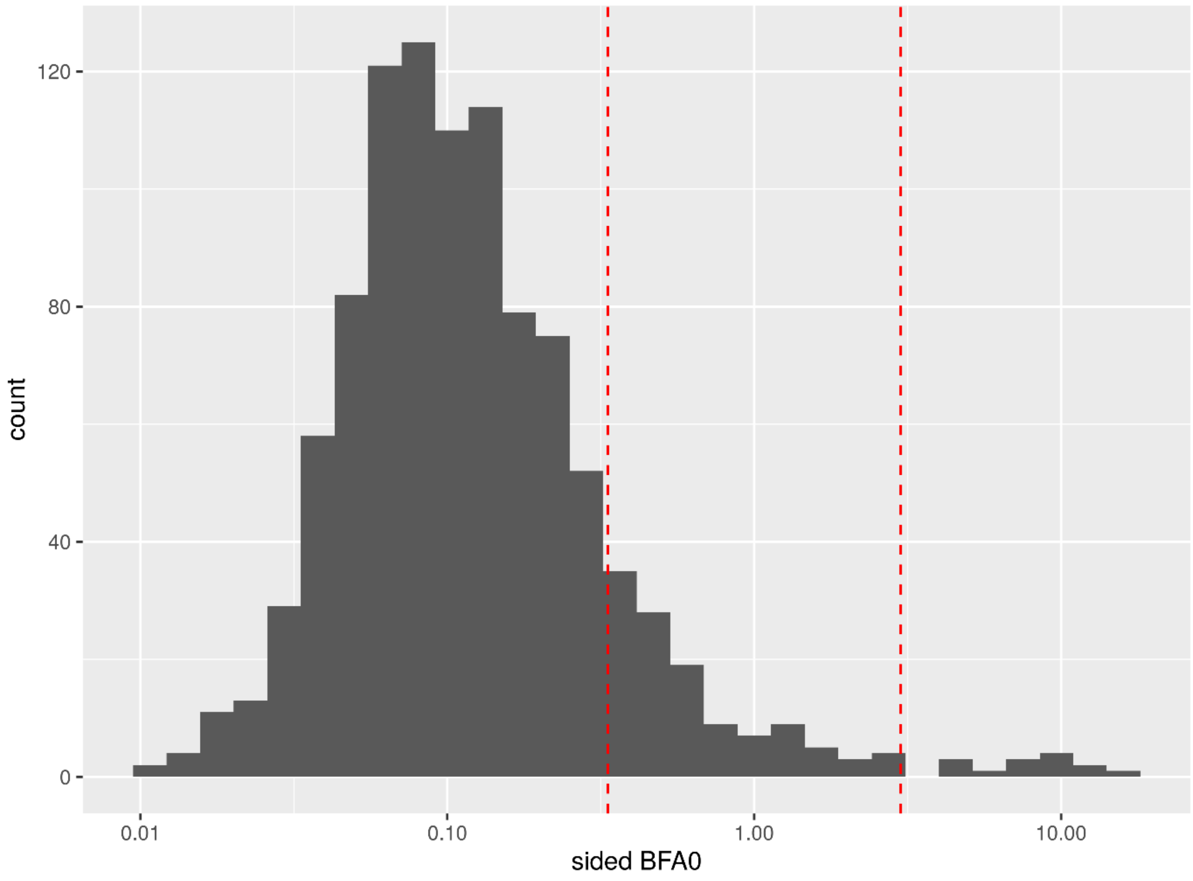


The red lines show the traditional thresholds at 1/3 and 3.

### Table S1

| fit index | 311 | ok? | 411a | ok? | 411b | ok? | 411c | ok? |
|---|---|---|---|---|---|---|---|---|
| chisq | 3.765 | | 0.842 | | 0.238 | | 0.160 | |
| df | 1.000 | | 1.000 | | 1.000 | | 1.000 | |
| p-value | 0.052 | good fit | 0.359 | good fit | 0.625 | good fit | 0.689 | good fit |
| chisq/df | 3.765 | unacceptable fit | 0.842 | good fit | 0.238 | good fit | 0.160 | good fit |
| rmsea | 0.042 | good fit | 0.000 | good fit | 0.000 | good fit | 0.000 | good fit |
| rmsea_lower | 0.000 | | 0.000 | | 0.000 | | 0.000 | |
| rmsea_upper | 0.091 | | 0.065 | | 0.053 | | 0.050 | |
| srmr | 0.019 | good fit | 0.003 | good fit | 0.001 | good fit | 0.001 | good fit |
| nnfi | 0.945 | unacceptable fit | 1.003 | unacceptable fit | 1.012 | unacceptable fit | 1.021 | unacceptable fit |
| cfi | 0.996 | good fit | 1.000 | good fit | 1.000 | good fit | 1.000 | good fit |

**Fit indices of mediation analyses of model 1.** chisq, chi squared; df, degrees of freedom
311: Indirect effect of social isolation on hippocampal volume via chronic stress
411a: Indirect effect of social isolation on executive functions via hippocampal volume
411b: Indirect effect of social isolation on memory via hippocampal volume
411c: Indirect effect of social isolation on processing speed via hippocampal volume

### Table S2

| **fit index** | **312** | **ok?** | **412a** | **ok?** | **412b** | **ok?** | **412c** | **ok?** |
|---|---|---|---|---|---|---|---|---|
| chisq | 9.260 | | 0.083 | | 0.958 | | 0.068 | |
| df | 5.000 | | 1.000 | | 1.000 | | 1.000 | |
| p-value | 0.099 | good fit | 0.773 | good fit | 0.328 | good fit | 0.794 | good fit |
| chisq/df | 1.852 | good fit | 0.083 | good fit | 0.958 | good fit | 0.068 | good fit |
| rmsea | 0.023 | good fit | 0.000 | good fit | 0.000 | good fit | 0.000 | good fit |
| rmsea_lower | 0.000 | | 0.000 | | 0.000 | | 0.000 | |
| rmsea_upper | 0.047 | | 0.045 | | 0.066 | | 0.043 | |
| srmr | 0.017 | good fit | 0.001 | good fit | 0.002 | good fit | 0.000 | good fit |
| nnfi | 0.972 | good fit | 1.014 | unacceptable fit | 1.001 | unacceptable fit | 1.026 | unacceptable fit |
| cfi | 0.994 | good fit | 1.000 | good fit | 1.000 | good fit | 1.000 | good fit |

**Fit indices of mediation analyses of model 2.** chisq, chi squared; df, degrees of freedom
312: Indirect effect of social isolation on hippocampal volume via chronic stress
412a: Indirect effect of social isolation on executive functions via hippocampal volume
412b: Indirect effect of social isolation on memory via hippocampal volume
412c: Indirect effect of social isolation on processing speed via hippocampal volume

#### Table S3

| dv | Model | Predictor | Estimate | 95% CI | p-value | FDR | BF |
|---|---|---|---|---|---|---|---|
| Hippo-campal Volume | 1 | LSNS_base | -5.500 | -9.122, -1.878 | 0.0015** | 0.0044** | 14.61** |
| | | LSNS_change | -4.894 | -8.492, -1.29 | 0.0039** | 0.0095** | 2.9 |
| | | age_base | -25.755 | -28.582, -22.929 | | | |
| | | age_change | -27.383 | -29.659, -25.115 | | | |
| | | gender | -48.683 | -85.261, -12.107 | | | |
| | 2 | LSNS_base | -5.672 | -9.503, -1.84 | 0.0019** | 0.0075** | 19.51** |
| | | LSNS_change | -4.928 | -8.741, -1.107 | 0.0058** | 0.0174* | 3.31* |
| | | age_base | -23.879 | -26.9, -20.858 | | | |
| | | age_change | -27.725 | -30.141, -25.32 | | | |
| | | gender | -47.733 | -85.365, -10.105 | | | |
| | | BMI | 18.831 | -0.946, 38.609 | | | |
| | | CESD | 13.369 | -5.716, 32.455 | | | |
| | | diabetes | -103.777 | -155.724, -51.827 | | | |
| | | education | -85.695 | -147.143, -24.244 | | | |
| | | hypertension | -29.051 | -69.373, 11.27 | | | |
| Executive Functions | 1 | LSNS_base | -0.026 | -0.035, -0.017 | 8.4e-09**** | 1.0e-07**** | 1.5e+06**** |
| | | LSNS_change | 0.003 | -0.011, 0.018 | 0.6787 | 0.787 | 0.08 |
| | | age_base | -0.020 | -0.027, -0.013 | | | |
| | | age_change | -0.053 | -0.063, -0.042 | | | |

| dv | Model | Predictor | Estimate | 95% CI | p-value | FDR | BF |
|---|---|---|---|---|---|---|---|
| | | gender | -0.074 | -0.166, 0.017 | | | |
| | 2 | LSNS_base | -0.015 | -0.025, -0.006 | 8e-04**** | 0.0046** | 43.65*** |
| | | LSNS_change | 0.006 | -0.009, 0.021 | 0.7842 | 0.8555 | 0.07 |
| | | age_base | -0.014 | -0.022, -0.007 | | | |
| | | age_change | -0.054 | -0.065, -0.043 | | | |
| | | gender | -0.121 | -0.214, -0.028 | | | |
| | | BMI | -0.079 | -0.128, -0.031 | | | |
| | | CESD | -0.137 | -0.183, -0.09 | | | |
| | | diabetes | -0.073 | -0.201, 0.054 | | | |
| | | education | -0.351 | -0.505, -0.196 | | | |
| | | hypertension | -0.078 | -0.177, 0.021 | | | |
| Memory | 1 | LSNS_base | -0.014 | -0.022, -0.006 | 5e-04**** | 0.002** | 49.05*** |
| | | LSNS_change | -0.013 | -0.026, 0 | 0.0262* | 0.0449* | 1.12 |
| | | age_base | -0.036 | -0.042, -0.029 | | | |
| | | age_change | -0.018 | -0.027, -0.009 | | | |
| | | gender | -0.381 | -0.465, -0.298 | | | |
| | 2 | LSNS_base | -0.008 | -0.016, 0.001 | 0.0452* | 0.0775 | 1.25 |
| | | LSNS_change | -0.009 | -0.023, 0.005 | 0.1046 | 0.1569 | 0.48 |
| | | age_base | -0.033 | -0.04, -0.026 | | | |
| | | age_change | -0.017 | -0.027, -0.008 | | | |

| dv | Model | Predictor | Estimate | 95% CI | p-value | FDR | BF |
|---|---|---|---|---|---|---|---|
| | | gender | -0.424 | -0.51, -0.338 | | | |
| | | BMI | -0.030 | -0.076, 0.015 | | | |
| | | CESD | -0.117 | -0.16, -0.073 | | | |
| | | diabetes | -0.045 | -0.162, 0.072 | | | |
| | | education | -0.166 | -0.306, -0.026 | | | |
| | | hypertension | 0.025 | -0.066, 0.116 | | | |
| Processing Speed | 1 | LSNS_base | -0.018 | -0.026, -0.011 | 1.7e-06**** | 1.0e-05**** | 9.4e+03**** |
| | | LSNS_change | -0.008 | -0.021, 0.005 | 0.1087 | 0.163 | 0.39 |
| | | age_base | -0.038 | -0.044, -0.032 | | | |
| | | age_change | -0.033 | -0.043, -0.024 | | | |
| | | gender | -0.112 | -0.188, -0.035 | | | |
| | 2 | LSNS_base | -0.018 | -0.026, -0.01 | 9.6e-06**** | 1e-04**** | 2.5e+03**** |
| | | LSNS_change | -0.012 | -0.025, 0.001 | 0.038* | 0.076 | 1.33 |
| | | age_base | -0.036 | -0.042, -0.029 | | | |
| | | age_change | -0.031 | -0.041, -0.022 | | | |
| | | gender | -0.135 | -0.214, -0.055 | | | |
| | | BMI | -0.025 | -0.066, 0.016 | | | |
| | | CESD | -0.024 | -0.063, 0.016 | | | |
| | | diabetes | 0.022 | -0.086, 0.131 | | | |
| | | education | -0.161 | -0.29, -0.031 | | | |

| dv | Model | Predictor | Estimate | 95% CI | p-value | FDR | BF |
|---|---|---|---|---|---|---|---|
| | | hypertension | -0.048 | -0.132, 0.036 | | | |

**Adjusted regression coefficients and measures of significance of models without interaction terms.** * p<0.05, BF>3; ** p<0.01, BF>10; *** p<0.001, BF>30; **** p<0.0001, BF>100; dv, dependent variable; CI, confidence interval; FDR, p-values after FDR-correction; BF, Bayes Factor in favour of alternative hypothesis; LSNS_base, baseline Lubben Social Network Score; LSNS_change, change in Lubben Social Network Score; CESD, Center for Epidemiological Studies-Depression
full model1: dv~LSNS_base+LSNS_change+age_base+age_change+gender
full model2: model1 + hypertension+diabetes+education+BMI+CESD
The unit of effect sizes on hippocampal volume and cognitive functions are $mm^3$/point on the LSNS and standard deviation/point on the LSNS, respectively.

#### Table S4

| dv | Model | Predictor | Estimate | 95% CI | p-value | FDR | BF |
|---|---|---|---|---|---|---|---|
| Hippo-campal Volume | 1 | LSNS_base*age_change | -0.556 | -1.099, -0.014 | 0.0223* | 0.0446* | 0.52 |
| | | LSNS_base | -5.033 | -8.682, -1.383 | | | |
| | | LSNS_change | -6.630 | -10.591, -2.665 | | | |
| | | age_base | -25.728 | -28.554, -22.902 | | | |
| | | age_change | -19.876 | -27.531, -12.217 | | | |
| | | gender | -48.216 | -84.786, -11.649 | | | |
| | 2 | LSNS_base*age_change | -0.538 | -1.107, 0.03 | 0.0318* | 0.076 | 0.63 |
| | | LSNS_base | -5.211 | -9.072, -1.35 | | | |
| | | LSNS_change | -6.541 | -10.702, -2.374 | | | |
| | | age_base | -23.854 | -26.874, -20.834 | | | |
| | | age_change | -20.416 | -28.492, -12.334 | | | |
| | | gender | -47.198 | -84.822, -9.579 | | | |
| | | BMI | 18.804 | -0.965, 38.576 | | | |
| | | CESD | 13.639 | -5.442, 32.721 | | | |
| | | diabetes | -103.725 | -155.653, -51.793 | | | |
| | | education | -85.668 | -147.094, -24.239 | | | |
| | | hypertension | -28.670 | -68.981, 11.639 | | | |
| Executive Functions | 1 | LSNS_base*age_change | 0.001 | -0.001, 0.003 | 0.7946 | 0.7946 | 0.06 |
| | | LSNS_base | -0.028 | -0.037, -0.018 | | | |
| | | LSNS_change | 0.006 | -0.01, 0.021 | | | |
| | | age_base | -0.020 | -0.027, -0.013 | | | |

| dv | Model | Predictor | Estimate | 95% CI | p-value | FDR | BF |
|---|---|---|---|---|---|---|---|
| | | age_change | -0.066 | -0.098, -0.033 | | | |
| | | gender | -0.075 | -0.166, 0.017 | | | |
| | 2 | LSNS_base*age_change | 0.002 | -0.001, 0.004 | 0.9062 | 0.9062 | 0.07 |
| | | LSNS_base | -0.018 | -0.028, -0.008 | | | |
| | | LSNS_change | 0.010 | -0.006, 0.026 | | | |
| | | age_base | -0.014 | -0.022, -0.007 | | | |
| | | age_change | -0.076 | -0.111, -0.041 | | | |
| | | gender | -0.122 | -0.215, -0.029 | | | |
| | | BMI | -0.079 | -0.127, -0.03 | | | |
| | | CESD | -0.137 | -0.184, -0.091 | | | |
| | | diabetes | -0.075 | -0.203, 0.053 | | | |
| | | education | -0.352 | -0.507, -0.197 | | | |
| | | hypertension | -0.080 | -0.179, 0.018 | | | |
| Memory | 1 | LSNS_base*age_change | 0.001 | -0.001, 0.003 | 0.7214 | 0.787 | 0.06 |
| | | LSNS_base | -0.015 | -0.024, -0.006 | | | |
| | | LSNS_change | -0.011 | -0.026, 0.003 | | | |
| | | age_base | -0.036 | -0.042, -0.029 | | | |
| | | age_change | -0.027 | -0.057, 0.004 | | | |
| | | gender | -0.382 | -0.465, -0.298 | | | |
| | 2 | LSNS_base*age_change | 0.001 | -0.001, 0.003 | 0.7451 | 0.8555 | 0.08 |
| | | LSNS_base | -0.009 | -0.018, 0.001 | | | |
| | | LSNS_change | -0.007 | -0.022, 0.008 | | | |

| dv | Model | Predictor | Estimate | 95% CI | p-value | FDR | BF |
|---|---|---|---|---|---|---|---|
| | | age_base | -0.033 | -0.04, -0.026 | | | |
| | | age_change | -0.028 | -0.059, 0.004 | | | |
| | | gender | -0.425 | -0.51, -0.339 | | | |
| | | BMI | -0.030 | -0.076, 0.015 | | | |
| | | CESD | -0.117 | -0.16, -0.074 | | | |
| | | diabetes | -0.046 | -0.163, 0.071 | | | |
| | | education | -0.167 | -0.307, -0.027 | | | |
| | | hypertension | 0.024 | -0.067, 0.116 | | | |
| Processing Speed | 1 | LSNS_base*age_change | -0.001 | -0.003, 0.001 | 0.17 | 0.2266 | 0.25 |
| | | LSNS_base | -0.017 | -0.025, -0.008 | | | |
| | | LSNS_change | -0.011 | -0.025, 0.003 | | | |
| | | age_base | -0.038 | -0.044, -0.032 | | | |
| | | age_change | -0.019 | -0.05, 0.011 | | | |
| | | gender | -0.111 | -0.187, -0.035 | | | |
| | 2 | LSNS_base*age_change | -0.001 | -0.003, 0.001 | 0.2411 | 0.3215 | 0.22 |
| | | LSNS_base | -0.017 | -0.025, -0.008 | | | |
| | | LSNS_change | -0.014 | -0.028, 0 | | | |
| | | age_base | -0.036 | -0.042, -0.029 | | | |
| | | age_change | -0.021 | -0.052, 0.011 | | | |
| | | gender | -0.134 | -0.213, -0.055 | | | |
| | | BMI | -0.025 | -0.066, 0.016 | | | |
| | | CESD | -0.023 | -0.063, 0.016 | | | |
| | | diabetes | 0.023 | -0.085, 0.132 | | | |
| | | education | -0.160 | -0.29, -0.031 | | | |

| dv | Model | Predictor | Estimate | 95% CI | p-value | FDR | BF |
|---|---|---|---|---|---|---|---|
| | | hypertension | -0.047 | -0.131, 0.037 | | | |

**Adjusted regression coefficients and measures of significance of models with interaction term of baseline social isolation with change in age.** * p<0.05, BF>3; dv, dependent variable; CI, confidence interval; FDR, p-values after FDR-correction; BF, Bayes Factor in favour of alternative hypothesis; LSNS_base, baseline Lubben Social Network Score; LSNS_change, change in Lubben Social Network Score; CESD, Center for Epidemiological Studies-Depression

full model1: dv~ LSNS_base*age_change+LSNS_base+LSNS_change+age_base+age_change+gender

full model2: model1 + hypertension+diabetes+education+BMI+CESD

#### Table S5

| dv | Model | Predictor | Estimate | 95% CI | p-value | BF |
|---|---|---|---|---|---|---|
| Hippocampal Volume | 1 | LSNS_base*LSNS_change | 0.11 | -0.61, 0.82 | 0.6146 | 0.03 |
| | | LSNS_base | -5.50 | -9.12, -1.88 | | |
| | | LSNS_change | -6.30 | -16.43, 3.82 | | |
| | | age_base | -25.75 | -28.58, -22.93 | | |
| | | age_change | -27.25 | -29.69, -24.82 | | |
| | | gender | -48.66 | -85.24, -12.09 | | |
| | 2 | LSNS_base*LSNS_change | 0.13 | -0.62, 0.88 | 0.6335 | 0.06 |
| | | LSNS_base | -5.67 | -9.5, -1.84 | | |
| | | LSNS_change | -6.67 | -17.4, 4.05 | | |
| | | age_base | -23.88 | -26.9, -20.86 | | |
| | | age_change | -27.57 | -30.14, -25.01 | | |
| | | gender | -47.73 | -85.36, -10.1 | | |
| | | BMI | 18.85 | -0.92, 38.63 | | |
| | | CESD | 13.34 | -5.74, 32.43 | | |
| | | diabetes | -103.63 | -155.58, -51.68 | | |
| | | education | -85.72 | -147.17, -24.27 | | |
| | | hypertension | -29.01 | -69.34, 11.3 | | |

**Adjusted regression coefficients and measures of significance of models with interaction term of baseline social isolation with change in social isolation.** dv, dependent variable; CI, confidence interval; BF, Bayes Factor in favour of alternative hypothesis; LSNS_base, baseline Lubben Social Network Score; LSNS_change, change in Lubben Social Network Score; CESD, Center for Epidemiological Studies-Depression
full model1: dv~LSNS_base+LSNS_change+age_base+age_change+gender
full model2: model1 + hypertension+diabetes+education+BMI+CESD

#### Table S6

| Mediator | dv | Model | Estimate | SE | z-value | p-value |
|---|---|---|---|---|---|---|
| TICS | Hippocampal Volume | 1 | -0.0005 | 0 | -0.56 | 0.29 |
| | | 2 | -0.0004 | 0 | -0.37 | 0.36 |
| Hippocampal Volume | Executive Functions | 1 | -0.0010 | 0 | -0.80 | 0.21 |
| | | 2 | -0.0013 | 0 | -0.94 | 0.17 |
| | Memory | 1 | -0.0010 | 0 | -0.82 | 0.20 |
| | | 2 | -0.0013 | 0 | -1.00 | 0.16 |
| | Processing Speed | 1 | -0.0002 | 0 | -0.27 | 0.40 |
| | | 2 | -0.0004 | 0 | -0.38 | 0.35 |

**Indirect effects of social isolation on hippocampal volume and cognitive functions.** dv, dependent variable; SE, standard error; TICS, Trierer Inventar zum chronischen Stress (stress questionnaire)
model1: corrected for baseline age, change in age and gender
model2: model1 + hypertension+diabetes+education+BMI+CESD

### Table S7

| dv | Model | Predictor | Estimate | 95% CI | p-value | FDR | BF |
|---|---|---|---|---|---|---|---|
| Hippocampal Volume | 1 | LSNS_base | -5.5 | -9.1, -1.9 | 0.0014** | 0.0042** | 18.65** |
| | | LSNS_change | -5.4 | -9, -1.8 | 0.0017** | 0.0042** | 7.6* |
| | | age_base | -25.7 | -28.6, -22.9 | | | |
| | | age_change | -25.5 | -28.3, -22.7 | | | |
| | | pandemic | -38.5 | -71.2, -5.8 | | | |
| | 2 | LSNS_base | -5.7 | -9.5, -1.9 | 0.0018** | 0.0073** | 20.97** |
| | | LSNS_change | -5.5 | -9.3, -1.7 | 0.0024** | 0.0073** | 6.8* |
| | | age_base | -23.9 | -26.9, -20.8 | | | |
| | | age_change | -25.8 | -28.8, -22.9 | | | |
| | | pandemic | -38.8 | -73.5, -3.8 | | | |

**Adjusted regression coefficients and measures of significance of hippocampal volume models adjusting for the effect of lockdown measures.** * p<0.05, BF>3; ** p<0.01, BF>10; *** p<0.001, BF>30; **** p<0.0001, BF>100; dv, dependent variable; CI, confidence interval; FDR, p-values after FDR-correction; BF, Bayes Factor in favour of alternative hypothesis; LSNS_base, baseline Lubben Social Network Score; LSNS_change, change in Lubben Social Network Score; CESD, Center for Epidemiological Studies-Depression; pandemic, 0/1 answered LSNS before/after beginning of 1st SARS-CoV-2 lockdown in Germany
full model1: dv~LSNS_base+LSNS_change+age_base+age_change+gender+pandemic
full model2: model1 + hypertension+diabetes+education+BMI+CESD
The effect sizes hardly change when including whether the LSNS was filled out after the begin of lockdown measures in the model. The effect of this control variable itself tends to be associated with smaller hippocampal volume but the confidence interval is very broad.

#### Table S8

| dv | Model | Predictor | Estimate | 95% CI | p-value | FDR | BF |
|---|---|---|---|---|---|---|---|
| Executive Functions | 1 | LSNS_base | -0.026 | -0.035, -0.017 | 7.7e-09**** | 9.2e-08**** | 1.7e+06**** |
| | | LSNS_change | 0.005 | -0.01, 0.019 | 0.733 | 0.7911 | 0.08 |
| | | age_base | -0.020 | -0.027, -0.013 | | | |
| | | age_change | -0.060 | -0.073, -0.048 | | | |
| | | pandemic | 0.133 | 0.004, 0.262 | | | |
| | 2 | LSNS_base | -0.015 | -0.025, -0.006 | 8e-04**** | 0.0046** | 36.51*** |
| | | LSNS_change | 0.007 | -0.008, 0.022 | 0.8314 | 0.9067 | 0.09 |
| | | age_base | -0.014 | -0.022, -0.007 | | | |
| | | age_change | -0.061 | -0.074, -0.048 | | | |
| | | pandemic | 0.136 | 0.001, 0.27 | | | |
| Memory | 1 | LSNS_base | -0.014 | -0.022, -0.006 | 5e-04**** | 0.0021** | 49.92*** |
| | | LSNS_change | -0.014 | -0.028, -0.001 | 0.0159* | 0.0272* | 1.89 |
| | | age_base | -0.036 | -0.042, -0.029 | | | |
| | | age_change | -0.009 | -0.02, 0.002 | | | |
| | | pandemic | -0.170 | -0.29, -0.05 | | | |
| | 2 | LSNS_base | -0.008 | -0.017, 0.001 | 0.0444* | 0.0761 | 1.33 |
| | | LSNS_change | -0.010 | -0.024, 0.003 | 0.0698 | 0.1047 | 0.85 |
| | | age_base | -0.033 | -0.04, -0.026 | | | |
| | | age_change | -0.010 | -0.021, 0.002 | | | |

| dv | Model | Predictor | Estimate | 95% CI | p-value | FDR | BF |
|---|---|---|---|---|---|---|---|
| | | pandemic | -0.158 | -0.283, -0.031 | | | |
| Processing Speed | 1 | LSNS_base | -0.018 | -0.026, -0.011 | 1.7e-06**** | 1.0e-05**** | 9.7e+03**** |
| | | LSNS_change | -0.008 | -0.021, 0.005 | 0.1055 | 0.1582 | 0.42 |
| | | age_base | -0.038 | -0.044, -0.032 | | | |
| | | age_change | -0.032 | -0.044, -0.021 | | | |
| | | pandemic | -0.020 | -0.136, 0.097 | | | |
| | 2 | LSNS_base | -0.018 | -0.026, -0.01 | 9.6e-06**** | 1e-04**** | 2.3e+03**** |
| | | LSNS_change | -0.012 | -0.025, 0.001 | 0.0366* | 0.0732 | 1.49 |
| | | age_base | -0.036 | -0.042, -0.029 | | | |
| | | age_change | -0.030 | -0.042, -0.018 | | | |
| | | pandemic | -0.020 | -0.14, 0.1 | | | |

**Adjusted regression coefficients and measures of significance of cognitive function models adjusting for the effect of lockdown measures.** * p<0.05, BF>3; ** p<0.01, BF>10; *** p<0.001, BF>30; **** p<0.0001, BF>100; dv, dependent variable; CI, confidence interval; FDR, p-values after FDR-correction; BF, Bayes Factor in favour of alternative hypothesis; LSNS_base, baseline Lubben Social Network Score; LSNS_change, change in Lubben Social Network Score; CESD, Center for Epidemiological Studies-Depression; pandemic, 0/1 answered LSNS before/after beginning of 1st SARS-CoV-2 lockdown in Germany

full model1: dv~LSNS_base+LSNS_change+age_base+age_change+gender+pandemic

full model2: model1 + hypertension+diabetes+education+BMI+CESD

The effect sizes hardly change when including whether the LSNS was filled out after the begin of lockdown measures in the model. The effect of this control variable itself is inconsistent between the different models.

#### Table S9

| dv | Model | Predictor | Estimate | 95% CI | p-value | FDR | BF |
|---|---|---|---|---|---|---|---|
| Hippo-campal Volume | 1 | LSNS_base | -3.9 | -7.3, -0.5 | 0.013* | 0.0222* | 2.39 |
| | | LSNS_change | -5.5 | -8.5, -2.4 | 2e-04**** | 7e-04**** | 32.58*** |
| | | age_base | -27.3 | -29.9, -24.6 | | | |
| | | age_change | -28.6 | -30.6, -26.5 | | | |
| | 2 | LSNS_base | -3.2 | -6.8, 0.4 | 0.0399* | 0.0684 | 0.97 |
| | | LSNS_change | -5.7 | -9, -2.5 | 3e-04**** | 0.0017** | 28.41** |
| | | age_base | -25.5 | -28.4, -22.7 | | | |
| | | age_change | -29.0 | -31.1, -26.8 | | | |

**Adjusted regression coefficients and measures of significance of hippocampal volume models based on datasets with reduced exclusion criteria.** * p<0.05, BF>3; ** p<0.01, BF>10; *** p<0.001, BF>30; **** p<0.0001, BF>100; dv, dependent variable; CI, confidence interval; FDR, p-values after FDR-correction; BF, Bayes Factor in favour of alternative hypothesis; LSNS_base, baseline Lubben Social Network Score; LSNS_change, change in Lubben Social Network Score; CESD, Center for Epidemiological Studies-Depression
full model1: dv~LSNS_base+LSNS_change+age_base+age_change+gender
full model2: model1 + hypertension+diabetes+education+BMI+CESD
Participants were not excluded for intake of cancer or centrally active medication and cognitive impairement
When applying less exclusion criteria, no major changes occur. For hippocampal volume baseline social isolation becomes deemphasized while the absolute effect size for change in social isolation becomes larger.

### Table S10

| dv | Model | Predictor | Estimate | 95% CI | p-value | FDR | BF |
|---|---|---|---|---|---|---|---|
| Executive Functions | 1 | LSNS_base | -0.030 | -0.038, -0.022 | 5.1e-13**** | 6.1e-12**** | 1.6e+10**** |
| | | LSNS_change | -0.009 | -0.021, 0.003 | 0.0759 | 0.1138 | 0.5 |
| | | age_base | -0.017 | -0.024, -0.011 | | | |
| | | age_change | -0.051 | -0.06, -0.042 | | | |
| | 2 | LSNS_base | -0.019 | -0.028, -0.011 | 4.5e-06**** | 5.4e-05**** | 4.6e+03**** |
| | | LSNS_change | -0.005 | -0.018, 0.008 | 0.2223 | 0.3335 | 0.27 |
| | | age_base | -0.011 | -0.018, -0.005 | | | |
| | | age_change | -0.052 | -0.062, -0.043 | | | |
| Memory | 1 | LSNS_base | -0.017 | -0.025, -0.009 | 2.6e-05**** | 1e-04**** | 745.27**** |
| | | LSNS_change | -0.015 | -0.027, -0.003 | 0.0079** | 0.0158* | 3.1* |
| | | age_base | -0.041 | -0.048, -0.033 | | | |
| | | age_change | -0.024 | -0.032, -0.015 | | | |
| | 2 | LSNS_base | -0.009 | -0.018, -0.001 | 0.0164* | 0.0328* | 2.91 |
| | | LSNS_change | -0.014 | -0.026, -0.001 | 0.0143* | 0.0328* | 2.49 |
| | | age_base | -0.038 | -0.045, -0.03 | | | |
| | | age_change | -0.025 | -0.034, -0.016 | | | |
| Processing Speed | 1 | LSNS_base | -0.015 | -0.022, -0.008 | 6.1e-06**** | 3.7e-05**** | 2.6e+03**** |
| | | LSNS_change | -0.016 | -0.026, -0.005 | 0.0022** | 0.0053** | 9.29* |
| | | age_base | -0.038 | -0.043, -0.033 | | | |

| dv | Model | Predictor | Estimate | 95% CI | p-value | FDR | BF |
|---|---|---|---|---|---|---|---|
| | | age_change | -0.035 | -0.043, -0.026 | | | |
| | 2 | LSNS_base | -0.012 | -0.019, -0.005 | 5e-04**** | 0.002** | 58.77*** |
| | | LSNS_change | -0.017 | -0.028, -0.006 | 0.0012** | 0.0037** | 21.76** |
| | | age_base | -0.035 | -0.04, -0.029 | | | |
| | | age_change | -0.033 | -0.041, -0.025 | | | |

**Adjusted regression coefficients and measures of significance of cognitive functions models based on datasets with reduced exclusion criteria.** * p<0.05, BF>3; ** p<0.01, BF>10; *** p<0.001, BF>30; **** p<0.0001, BF>100; dv, dependent variable; CI, confidence interval; FDR, p-values after FDR-correction; BF, Bayes Factor in favour of alternative hypothesis; LSNS_base, baseline Lubben Social Network Score; LSNS_change, change in Lubben Social Network Score; CESD, Center for Epidemiological Studies-Depression
full model1: dv~LSNS_base+LSNS_change+age_base+age_change+gender
full model2: model1 + hypertension+diabetes+education+BMI+CESD
When applying less exclusion criteria, the direction and magnitude of effect sizes tends to stay the same. The direction of change in social isolation becomes negative but is still small. Most significances are more pronounced. Given the larger sample size, this is to be expected.

### Table S11

| dv | Model | Predictor | Estimate | 95% CI | p-value | FDR | BF |
|---|---|---|---|---|---|---|---|
| Hippo-campal Volume | 1 | mean LSNS | -6.9 | -11.3, -2.6 | 9e-04**** | 0.0036** | 26.01** |
| | | LSNS within | -4.7 | -8.3, -1.1 | 0.0054** | 0.0161* | 1.92 |
| | | mean age | -26.1 | -29.4, -22.7 | | | |
| | | age within | -26.5 | -28.8, -24.2 | | | |
| | 2 | mean LSNS | -6.7 | -11.2, -2.1 | 0.0021** | 0.0101* | 17.76** |
| | | LSNS within | -4.6 | -8.4, -0.8 | 0.009** | 0.027* | 1.87 |
| | | mean age | -24.6 | -28.1, -21 | | | |
| | | age within | -26.8 | -29.2, -24.4 | | | |

**Adjusted regression coefficients and measures of significance of hippocampal volume models only including participants with two timepoints.** * p<0.05, BF>3; ** p<0.01, BF>10; *** p<0.001, BF>30; **** p<0.0001, BF>100; dv, dependent variable; CI, confidence interval; FDR, p-values after FDR-correction; BF, Bayes Factor in favour of alternative hypothesis; mean LSNS, subject's mean Lubben Social Network Score; LSNS within, within subject variation in Lubben Social Network Score; mean age, subject's mean age; age within, within subject variation in age; CESD, Center for Epidemiological Studies-Depression
full model1: dv~LSNS_base+LSNS_change+age_base+age_change+gender
full model2: model1 + hypertension+diabetes+education+BMI+CESD
In this sensitivity analysis only participants with two timepoints were included and standard mean and within scores rather than baseline and change scores were calculated. In terms of effect size and direction our original model is corroborated. Smaller measures of significance in this smaller sample were expectable.

#### Table S12

| dv | Model | Predictor | Estimate | 95% CI | p-value | FDR | BF |
|---|---|---|---|---|---|---|---|
| Executive Functions | 1 | mean LSNS | -0.027 | -0.037, -0.016 | 5.9e-07**** | 7.1e-06**** | 2.7e+04**** |
| | | LSNS within | 0.005 | -0.011, 0.021 | 0.7316 | 0.7607 | 0.08 |
| | | mean age | -0.014 | -0.023, -0.006 | | | |
| | | age within | -0.055 | -0.066, -0.045 | | | |
| | 2 | mean LSNS | -0.016 | -0.027, -0.005 | 0.0025** | 0.0101* | 16.1** |
| | | LSNS within | 0.005 | -0.012, 0.021 | 0.7176 | 0.7829 | 0.11 |
| | | mean age | -0.008 | -0.016, 0.001 | | | |
| | | age within | -0.055 | -0.065, -0.044 | | | |
| Memory | 1 | mean LSNS | -0.010 | -0.019, 0 | 0.0225* | 0.045* | 2.02 |
| | | LSNS within | -0.010 | -0.024, 0.004 | 0.0874 | 0.1498 | 0.43 |
| | | mean age | -0.031 | -0.039, -0.024 | | | |
| | | age within | -0.018 | -0.027, -0.009 | | | |
| | 2 | mean LSNS | -0.006 | -0.016, 0.004 | 0.1243 | 0.2131 | 0.66 |
| | | LSNS within | -0.006 | -0.021, 0.008 | 0.2046 | 0.307 | 0.3 |
| | | mean age | -0.027 | -0.035, -0.02 | | | |
| | | age within | -0.016 | -0.025, -0.006 | | | |
| Processing Speed | 1 | mean LSNS | -0.015 | -0.024, -0.006 | 4e-04**** | 0.0027** | 58.1*** |
| | | LSNS within | -0.006 | -0.02, 0.009 | 0.2218 | 0.3305 | 0.2 |
| | | mean age | -0.039 | -0.046, -0.032 | | | |
| | | age within | -0.033 | -0.043, -0.023 | | | |

| dv | Model | Predictor | Estimate | 95% CI | p-value | FDR | BF |
|---|---|---|---|---|---|---|---|
| | 2 | mean LSNS | -0.014 | -0.024, -0.005 | 0.0017** | 0.0101* | 21.84** |
| | | LSNS within | -0.011 | -0.026, 0.004 | 0.0721 | 0.1441 | 0.66 |
| | | mean age | -0.038 | -0.045, -0.031 | | | |
| | | age within | -0.031 | -0.041, -0.021 | | | |

**Adjusted regression coefficients and measures of significance of cognitive functions models only including participants with two timepoints.** * p<0.05, BF>3; ** p<0.01, BF>10; *** p<0.001, BF>30; **** p<0.0001, BF>100; dv, dependent variable; CI, confidence interval; FDR, p-values after FDR-correction; BF, Bayes Factor in favour of alternative hypothesis; mean LSNS, subject's mean Lubben Social Network Score; LSNS within, within subject variation in Lubben Social Network Score; mean age, subject's mean age; age within, within subject variation in age; CESD, Center for Epidemiological Studies-Depression
full model1: dv~LSNS_base+LSNS_change+age_base+age_change+gender
full model2: model1 + hypertension+diabetes+education+BMI+CESD
In this sensitivity analysis only participants with two timepoints were included and standard mean and within scores rather than baseline and change scores were calculated. As for hippocampal volume, this sensitivity analysis corroborates our original model.

### Table S13

| dv | Model | Predictor | Estimate | 95% CI | p-value | FDR | BF |
|---|---|---|---|---|---|---|---|
| Hippocampal Volume | 1 | LSNS_base | -5.5 | -9.1, -1.9 | 0.0015** | 0.0044** | 19.53** |
| | | LSNS_change | -4.9 | -8.5, -1.3 | 0.0038** | 0.0091** | 2.34 |
| | | age_base | -25.8 | -28.6, -22.9 | | | |
| | | age_change | -27.4 | -29.6, -25.1 | | | |
| | 2 | LSNS_base | -5.7 | -9.5, -1.9 | 0.0018** | 0.0073** | 17.34** |
| | | LSNS_change | -4.9 | -8.7, -1.1 | 0.0055** | 0.0164* | 3.37* |
| | | age_base | -24.2 | -27.2, -21.1 | | | |
| | | age_change | -27.7 | -30.1, -25.3 | | | |
| | | hypertension | -15.6 | -57.1, 25.8 | | | |

**Adjusted regression coefficients and measures of significance of hippocampal volume models using a hypertension cut-off of 140mmHg.** * p<0.05, BF>3; ** p<0.01, BF>10; *** p<0.001, BF>30; **** p<0.0001, BF>100; dv, dependent variable; CI, confidence interval; FDR, p-values after FDR-correction; BF, Bayes Factor in favour of alternative hypothesis; mean LSNS, subject's mean Lubben Social Network Score; LSNS within, within subject variation in Lubben Social Network Score; mean age, subject's mean age; age within, within subject variation in age; CESD, Center for Epidemiological Studies-Depression
full model1: dv~LSNS_base+LSNS_change+age_base+age_change+gender
full model2: model1 + hypertension+diabetes+education+BMI+CESD
Social isolation significantly predicts hippocampal volume after multiplicity control. Bayes Factors provide strong evidence in favour of the alternative hypotheses for baseline social isolation and anecdotal to moderate evidence for change in social isolation. The effect size of one point on the LSNS is equivalent to a baseline age difference of around two and a half months.

### Table S14

| dv | Model | Predictor | Estimate | 95% CI | p-value | FDR | BF |
|---|---|---|---|---|---|---|---|
| Executive Functions | 1 | LSNS_base | -0.026 | -0.035, -0.017 | 8.2e-09 | 9.9e-08 | 1.5e+06 |
| | | LSNS_change | 0.003 | -0.011, 0.018 | 0.6775 | 0.7893 | 0.08 |
| | | age_base | -0.019 | -0.026, -0.012 | | | |
| | | age_change | -0.053 | -0.063, -0.042 | | | |
| | 2 | LSNS_base | -0.015 | -0.025, -0.006 | 8e-04 | 0.0047 | 50.05 |
| | | LSNS_change | 0.006 | -0.009, 0.021 | 0.78 | 0.851 | 0.09 |
| | | age_base | -0.013 | -0.021, -0.006 | | | |
| | | age_change | -0.054 | -0.065, -0.044 | | | |
| | | hypertension | -0.120 | -0.222, -0.018 | | | |
| Memory | 1 | LSNS_base | -0.014 | -0.022, -0.006 | 5e-04 | 0.002 | 48.6 |
| | | LSNS_change | -0.013 | -0.026, 0 | 0.0265 | 0.0454 | 1.11 |
| | | age_base | -0.036 | -0.042, -0.029 | | | |
| | | age_change | -0.018 | -0.027, -0.009 | | | |
| | 2 | LSNS_base | -0.007 | -0.016, 0.001 | 0.0501 | 0.086 | 1.15 |
| | | LSNS_change | -0.009 | -0.023, 0.005 | 0.1033 | 0.1549 | 0.49 |
| | | age_base | -0.032 | -0.039, -0.025 | | | |
| | | age_change | -0.018 | -0.027, -0.008 | | | |
| | | hypertension | -0.006 | -0.1, 0.089 | | | |
| Processing Speed | 1 | LSNS_base | -0.018 | -0.025, -0.01 | 2.4e-06 | 1.4e-05 | 6.8e+03 |
| | | LSNS_change | -0.008 | -0.021, 0.005 | 0.1074 | 0.1611 | 0.36 |
| | | age_base | -0.038 | -0.044, -0.032 | | | |
| | | age_change | -0.034 | -0.043, -0.024 | | | |

| dv | Model | Predictor | Estimate | 95% CI | p-value | FDR | BF |
|---|---|---|---|---|---|---|---|
| | | LSNS_base | -0.018 | -0.026, -0.009 | 1.2e-05 | 1e-04 | 1.8e+03 |
| | | LSNS_change | -0.012 | -0.025, 0.001 | 0.0371 | 0.0741 | 1.56 |
| | 2 | age_base | -0.037 | -0.043, -0.031 | | | |
| | | age_change | -0.032 | -0.041, -0.022 | | | |
| | | hypertension | -0.002 | -0.088, 0.085 | | | |

**Adjusted regression coefficients and measures of significance of cognitive functions models using a hypertension cut-off of 140mmHg.** * p<0.05, BF>3; ** p<0.01, BF>10; *** p<0.001, BF>30; **** p<0.0001, BF>100; dv, dependent variable; CI, confidence interval; FDR, p-values after FDR-correction; BF, Bayes Factor in favour of alternative hypothesis; mean LSNS, subject's mean Lubben Social Network Score; LSNS within, within subject variation in Lubben Social Network Score; mean age, subject's mean age; age within, within subject variation in age; CESD, Center for Epidemiological Studies-Depression
full model1: dv~LSNS_base+LSNS_change+age_base+age_change+gender
full model2: model1 + hypertension+diabetes+education+BMI+CESD
Baseline social isolation significantly predicts cognitive functions after FDR-correction and BFs provide very strong to decisive evidence in favour of the alternative hypotheses. Only for model 2 of memory evidence is weak. No association of change in social isolation with executive functions is detected and evidence for associations with memory and processing speed are limited.

#### Table S15

| dv | Model | Predictor | Estimate | 95% CI | p-value | FDR | BF |
|---|---|---|---|---|---|---|---|
| Hippo-campal Volume | 1 | LSNS_base | -7.3 | -11.2, -3.4 | 1e-04 | 4e-04 | 192.27 |
| | | LSNS_change | -4.5 | -8.2, -0.8 | 0.0093 | 0.0223 | 1.18 |
| | | age_base | -24.5 | -27.5, -21.6 | | | |
| | | age_change | -27.7 | -30.1, -25.3 | | | |
| | 2 | LSNS_base | -7.1 | -11.2, -3 | 4e-04 | 0.0042 | 81.34 |
| | | LSNS_change | -4.6 | -8.6, -0.7 | 0.0103 | 0.0309 | 1.7 |
| | | age_base | -22.4 | -25.6, -19.2 | | | |
| | | age_change | -27.7 | -30.2, -25.2 | | | |

**Adjusted regression coefficients and measures of significance of hippocampal volume models excluding participants with MMSE score < 27.** * p<0.05, BF>3; ** p<0.01, BF>10; *** p<0.001, BF>30; **** p<0.0001, BF>100; dv, dependent variable; CI, confidence interval; FDR, p-values after FDR-correction; BF, Bayes Factor in favour of alternative hypothesis; mean LSNS, subject's mean Lubben Social Network Score; LSNS within, within subject variation in Lubben Social Network Score; mean age, subject's mean age; age within, within subject variation in age; CESD, Center for Epidemiological Studies-Depression
full model1: dv~LSNS_base+LSNS_change+age_base+age_change+gender
full model2: model1 + hypertension+diabetes+education+BMI+CESD
Social isolation significantly predicts hippocampal volume after multiplicity control. Bayes Factors provide strong evidence in favour of the alternative hypotheses for baseline social isolation and anecdotal to moderate evidence for change in social isolation. The effect size of one point on the LSNS is equivalent to a baseline age difference of around two and a half months.

#### Table S16

| Model | Predictor | Estimate | 95% CI | p-value | FDR | BF |
|---|---|---|---|---|---|---|
| 1 | LSNS_base | -0.023 | -0.033, -0.013 | 1.9e-06 | 2.3e-05 | 9.4e+03 |
| | LSNS_change | 0.004 | -0.01, 0.019 | 0.7159 | 0.7159 | 0.08 |
| | age_base | -0.016 | -0.024, -0.009 | | | |
| | age_change | -0.058 | -0.068, -0.047 | | | |
| 2 | LSNS_base | -0.014 | -0.024, -0.003 | 0.0049 | 0.0194 | 8.81 |
| | LSNS_change | 0.007 | -0.008, 0.022 | 0.8175 | 0.8384 | 0.09 |
| | age_base | -0.012 | -0.02, -0.004 | | | |
| | age_change | -0.058 | -0.069, -0.048 | | | |
| 1 | LSNS_base | -0.014 | -0.023, -0.005 | 0.0011 | 0.0034 | 24.53 |
| | LSNS_change | -0.013 | -0.027, 0.001 | 0.0308 | 0.0615 | 1.08 |
| | age_base | -0.033 | -0.039, -0.026 | | | |
| | age_change | -0.028 | -0.038, -0.018 | | | |
| 2 | LSNS_base | -0.009 | -0.018, 0.001 | 0.0355 | 0.0852 | 1.66 |
| | LSNS_change | -0.009 | -0.023, 0.006 | 0.1181 | 0.169 | 0.51 |
| | age_base | -0.029 | -0.036, -0.021 | | | |
| | age_change | -0.026 | -0.036, -0.016 | | | |
| 1 | LSNS_base | -0.016 | -0.024, -0.008 | 1e-04 | 4e-04 | 198.61 |
| | LSNS_change | -0.007 | -0.02, 0.006 | 0.1509 | 0.2012 | 0.31 |
| | age_base | -0.038 | -0.045, -0.032 | | | |
| | age_change | -0.038 | -0.047, -0.028 | | | |
| 2 | LSNS_base | -0.014 | -0.023, -0.005 | 8e-04 | 0.005 | 40.5 |
| | LSNS_change | -0.010 | -0.023, 0.004 | 0.0827 | 0.1418 | 0.8 |
| | age_base | -0.036 | -0.043, -0.029 | | | |
| | age_change | -0.035 | -0.045, -0.025 | | | |

**Adjusted regression coefficients and measures of significance of cognitive functions models excluding participants with MMSE score < 27.** * p<0.05, BF>3; ** p<0.01, BF>10; *** p<0.001, BF>30; **** p<0.0001, BF>100; dv, dependent variable; CI, confidence interval; FDR, p-values after FDR-correction; BF, Bayes Factor in favour of alternative hypothesis; mean LSNS, subject's mean Lubben Social Network Score; LSNS within, within subject variation in Lubben Social Network Score; mean age, subject's mean age; age within, within subject variation in age; CESD, Center for Epidemiological Studies-Depression
full model1: dv~LSNS_base+LSNS_change+age_base+age_change+gender
full model2: model1 + hypertension+diabetes+education+BMI+CESD
Baseline social isolation significantly predicts cognitive functions after FDR-correction and BFs provide very strong to decisive evidence in favour of the alternative hypotheses. Only for model 2 of memory evidence is weak. No association of change in social isolation with executive functions is detected and evidence for associations with memory and processing speed are limited.

**Table S17**

| dv | Model | gender | Predictor | Estimate | 95% CI | p-value | FDR |
|---|---|---|---|---|---|---|---|
| Hippo-campal Volume | 1 | female | LSNS_base | -7.265 | -12.546, -1.984 | 0.0036** | 0.0142* |
| | | | LSNS_change | -3.826 | -8.389, 0.75 | 0.0503 | 0.1006 |
| | | | LSNS_base*age_change | -0.311 | -0.992, 0.37 | 0.1847 | 0.2463 |
| | | | LSNS_base*LSNS_change | -0.026 | -0.865, 0.812 | 0.4755 | |
| | 1 | male | LSNS_base | -4.418 | -9.407, 0.572 | 0.0414* | 0.0827 |
| | | | LSNS_change | -5.821 | -11.462, -0.17 | 0.0218* | 0.0655 |
| | | | LSNS_base*age_change | -0.793 | -1.656, 0.066 | 0.0356* | 0.0827 |
| | | | LSNS_base*LSNS_change | 0.426 | -0.831, 1.696 | 0.7466 | |
| | 2 | female | LSNS_base | -9.402 | -15.042, -3.762 | 6e-04**** | 0.0068** |
| | | | LSNS_change | -3.452 | -8.28, 1.395 | 0.0807 | 0.1614 |
| | | | LSNS_base*age_change | -0.255 | -0.971, 0.462 | 0.2422 | 0.3229 |
| | | | LSNS_base*LSNS_change | 0.027 | -0.842, 0.895 | 0.5248 | |
| | 2 | male | LSNS_base | -3.046 | -8.299, 2.207 | 0.1277 | 0.2554 |
| | | | LSNS_change | -6.344 | -12.289, -0.39 | 0.0185* | 0.1111 |
| | | | LSNS_base*age_change | -0.796 | -1.692, 0.095 | 0.0403* | 0.1209 |
| | | | LSNS_base*LSNS_change | 0.448 | -0.876, 1.783 | 0.7464 | |
| Executive Functions | 1 | female | LSNS_base | -0.032 | -0.045, -0.018 | 1.6e-06**** | 1.9e-05**** |
| | | | LSNS_change | -0.006 | -0.026, 0.014 | 0.2797 | 0.3357 |

| dv | Model | gender | Predictor | Estimate | 95% CI | p-value | FDR |
|---|---|---|---|---|---|---|---|
| | | | LSNS_base*age_change | 0.001 | -0.002, 0.004 | 0.7135 | 0.7135 |
| | 1 | male | LSNS_base | -0.022 | -0.034, -0.009 | 4e-04**** | 0.0022** |
| | | | LSNS_change | 0.013 | -0.007, 0.033 | 0.9021 | 0.9021 |
| | | | LSNS_base*age_change | 0.001 | -0.002, 0.005 | 0.8056 | 0.8789 |
| | 2 | female | LSNS_base | -0.020 | -0.034, -0.006 | 0.0032** | 0.019* |
| | | | LSNS_change | 0.001 | -0.02, 0.022 | 0.547 | 0.6564 |
| | | | LSNS_base*age_change | 0.002 | -0.002, 0.005 | 0.8642 | 0.8642 |
| | 2 | male | LSNS_base | -0.012 | -0.025, 0 | 0.0293* | 0.1173 |
| | | | LSNS_change | 0.012 | -0.009, 0.033 | 0.8653 | 0.8653 |
| | | | LSNS_base*age_change | 0.002 | -0.002, 0.005 | 0.8482 | 0.8653 |
| Memory | 1 | female | LSNS_base | -0.011 | -0.023, 0.001 | 0.0345* | 0.0827 |
| | | | LSNS_change | -0.017 | -0.034, -0.001 | 0.0218* | 0.0655 |
| | | | LSNS_base*age_change | 0.000 | -0.003, 0.003 | 0.5141 | 0.5609 |
| | 1 | male | LSNS_base | -0.016 | -0.028, -0.004 | 0.0035** | 0.0141* |
| | | | LSNS_change | -0.007 | -0.028, 0.013 | 0.2454 | 0.4081 |
| | | | LSNS_base*age_change | 0.001 | -0.002, 0.005 | 0.7892 | 0.8789 |
| | 2 | female | LSNS_base | -0.004 | -0.017, 0.008 | 0.2417 | 0.3229 |
| | | | LSNS_change | -0.015 | -0.032, 0.003 | 0.0494* | 0.1185 |
| | | | LSNS_base*age_change | 0.000 | -0.002, 0.003 | 0.612 | 0.6677 |
| | 2 | male | LSNS_base | -0.010 | -0.022, 0.003 | 0.0644 | 0.1544 |

| dv | Model | gender | Predictor | Estimate | 95% CI | p-value | FDR |
|---|---|---|---|---|---|---|---|
| | | | LSNS_change | -0.002 | -0.023, 0.02 | 0.4446 | 0.5928 |
| | | | LSNS_base*age_change | 0.001 | -0.002, 0.004 | 0.7429 | 0.8653 |
| Processing Speed | 1 | female | LSNS_base | -0.017 | -0.028, -0.005 | 0.0028** | 0.0142* |
| | | | LSNS_change | -0.009 | -0.026, 0.009 | 0.1632 | 0.2448 |
| | | | LSNS_base*age_change | -0.002 | -0.005, 0.001 | 0.127 | 0.2177 |
| | 1 | male | LSNS_base | -0.020 | -0.03, -0.01 | 6.6e-05**** | 8e-04**** |
| | | | LSNS_change | -0.006 | -0.025, 0.013 | 0.2721 | 0.4081 |
| | | | LSNS_base*age_change | 0.000 | -0.003, 0.003 | 0.4427 | 0.5903 |
| | 2 | female | LSNS_base | -0.016 | -0.029, -0.004 | 0.0053** | 0.0211* |
| | | | LSNS_change | -0.015 | -0.033, 0.002 | 0.0449* | 0.1185 |
| | | | LSNS_base*age_change | -0.001 | -0.004, 0.002 | 0.1922 | 0.3229 |
| | 2 | male | LSNS_base | -0.018 | -0.029, -0.008 | 4e-04**** | 0.0051** |
| | | | LSNS_change | -0.007 | -0.028, 0.013 | 0.2368 | 0.4059 |
| | | | LSNS_base*age_change | 0.000 | -0.003, 0.003 | 0.4265 | 0.5928 |

**Adjusted regression coefficients and measures of significance of models stratified by gender**. * p<0.05 ; ** p<0.01; *** p<0.001; **** p<0.0001; dv, dependent variable; CI, confidence interval; FDR, p-values after FDR-correction; LSNS_base, baseline Lubben Social Network Score; LSNS_change, change in Lubben Social Network Score; CESD, Center for Epidemiological Studies-Depression
full model1: dv~LSNS_base+LSNS_change+age_base+age_change
full model2: model1 + hypertension+diabetes+education+BMI+CESD
The unit of effect sizes on hippocampal volume and cognitive functions for non-interaction models are $mm^3$/point on the LSNS and standard deviation/point on the LSNS, respectively. For interaction models the unit in the denominator is multiplied by year or point on the LSNS.

### Table S18

| dv | predictor | estimate | se | p-value | q value |
|---|---|---|---|---|---|
| ΔHCV | LSNS_base | -0.002 | 0.005 | 0.315 | 0.420 |
| ΔLSNS | HCV_base | -0.139 | 0.175 | 0.213 | 0.284 |
| ΔEF | LSNS_base | -0.014 | 0.007 | 0.029* | 0.116 |
| ΔLSNS | EF_base | -0.149 | 0.170 | 0.189 | 0.284 |
| ΔMemo | LSNS_base | 0.001 | 0.006 | 0.576 | 0.576 |
| ΔLSNS | Memo_base | -0.308 | 0.168 | 0.033* | 0.133 |
| ΔPS | LSNS_base | -0.005 | 0.008 | 0.250 | 0.420 |
| ΔLSNS | PS_base | -0.102 | 0.179 | 0.285 | 0.285 |

**Relevant Regressions of Bivariate Latent Change Score Models.** *, $p < 0.05$; dv, dependent variable; se, standard error; _base, baseline score of; Δ, change in; LSNS, Lubben Social Network Score; HCV, z-transformed hippocampal volume; EF, executive functions; Memo, memory; PS, processing speed

#### Table S19

| BFA0 | FWER in % | n |
|---:|---:|---:|
| 15.744 | 1.18 | 1 |
| 13.634 | 2.36 | 2 |
| 13.139 | 3.51 | 3 |
| 10.926 | 4.66 | 4 |
| 10.632 | 5.79 | 5 |
| 9.196 | 6.91 | 6 |
| 8.728 | 8.02 | 7 |
| 8.510 | 9.12 | 8 |
| 7.749 | 10.20 | 9 |
| 7.191 | 11.28 | 10 |
| 6.081 | 12.34 | 11 |
| 4.746 | 13.39 | 12 |
| 4.044 | 14.42 | 13 |
| 4.003 | 15.45 | 14 |

**Simulated Bayes Factors above the threshold of 3.** BFA0, Sided Bayes factor in favour of the alternative hypothesis; FWER, familywise error rate if the threshold would be set just below BFA0
In the simulation with randomly simulated values for our predictors of interest, 14 BFs exceeded the standard threshold of three. Given a family size of 12 tests, a threshold of 10.75 would maintain the FWER below 5%.

### Table S20

| Category | BFA0b > 3 in % | 3 >= BFA0b >= 1/3 in % | BFA0b < 1/3 in % | BFA0c > 3 in % | 3 >= BFA0c >= 1/3 in % | BFA0c < 1/3 in % | n |
|---|---|---|---|---|---|---|---|
| overall | 44.23 | 31.41 | 24.36 | 28.85 | 30.45 | 40.71 | 312 |
| model 1 | 45.51 | 30.13 | 24.36 | 30.13 | 30.13 | 39.74 | 156 |
| model 2 | 42.95 | 32.69 | 24.36 | 27.56 | 30.77 | 41.67 | 156 |
| effect = 0.1 | 9.62 | 38.46 | 51.92 | 5.77 | 24.04 | 70.19 | 104 |
| effect = 0.2 | 37.50 | 44.23 | 18.27 | 21.15 | 39.42 | 39.42 | 104 |
| effect = 0.5 | 85.58 | 11.54 | 2.88 | 59.62 | 27.88 | 12.50 | 104 |

**Results of Power Simulation of Bayes Factors.** BFA0b, Sided Bayes factor in favour of the alternative hypothesis of baseline social isolation; BFA0c, Sided Bayes factor in favour of the alternative hypothesis of change in social isolation; n, number of simulations in the category; model 1, model with reduced number of control variables; model 2, model with full number of control variables; effect, effect size per point in the Lubben Social Network Scale in years of baseline age
Percentages of Bayes Factors giving moderate or stronger evidence in favour of the alternative hypothesis (>3), giving anecdotal evidence (3>=BF>=1/3) and giving moderate or stronger evidence in favour of the null hypothesis (< 1/3).

### Table S21

| Category | BFA0b > 10.75 in % | 10.75 >= BFA0b >= 1/3 in % | BFA0b < 1/3 in % | BFA0c > 10.75 in % | 10.75 >= BFA0c >= 1/3 in % | BFA0c < 1/3 in % | n |
|---|---|---|---|---|---|---|---|
| overall | 37.18 | 38.46 | 24.36 | 20.83 | 38.46 | 40.71 | 312 |
| model 1 | 38.46 | 37.18 | 24.36 | 21.79 | 38.46 | 39.74 | 156 |
| model 2 | 35.90 | 39.74 | 24.36 | 19.87 | 38.46 | 41.67 | 156 |
| effect = 0.1 | 5.77 | 42.31 | 51.92 | 0.96 | 28.85 | 70.19 | 104 |
| effect = 0.2 | 24.04 | 57.69 | 18.27 | 14.42 | 46.15 | 39.42 | 104 |
| effect = 0.5 | 81.73 | 15.38 | 2.88 | 47.12 | 40.38 | 12.50 | 104 |

**Results of Power Simulation of Bayes Factors with adjusted thresholds for a family of 12 tests.** BFA0b, Sided Bayes factor in favour of the alternative hypothesis of baseline social isolation; BFA0c, Sided Bayes factor in favour of the alternative hypothesis of change in social isolation; n, number of simulations in the category; model 1, model with reduced number of control variables; model 2, model with full number of control variables; effect, effect size per point in the Lubben Social Network Scale in years of baseline age. Percentages of Bayes Factors giving moderate or stronger evidence in favour of the alternative hypothesis (>10.75), giving anecdotal evidence (10.75>=BF>=1/3) and giving moderate or stronger evidence in favour of the null hypothesis (< 1/3).
